# Oestrogen and autoimmune disease activity: A scoping review

**DOI:** 10.1101/2025.08.12.25333495

**Authors:** Lily Summerhayes, Kayleigh E Easey

**Author notes:** Corresponding author: Kayleigh Easey.

## Abstract

**Introduction:** Symptoms of autoimmune disorders (ADs) have been observed to fluctuate around reproductive events, likely due to hormone changes during these timepoints. There is conflicting evidence for how changes in oestrogen levels may be related to a reduction of AD symptoms, and how this varies across specific ADs.

**Methods:** A scoping review was conducted according to PRISMA guidelines, to harmonise the current evidence within this field. PubMed was searched and data extracted based on AD type (rheumatoid arthritis (RA), multiple sclerosis (MS) and type-one diabetes (T1D)), AD symptom measured, oestrogen hormone type, demographics (population location, age, gender, human/animal study), and summarised.

**Results:** A total of 68 studies were included; 35 measured RA, 25 for MS, 7 for T1D, and 1 for both RA and MS combined. Most studies used a female only sample (74%). Of the 68 included studies, 34 showed oestrogen hormones were associated with improvements in AD symptoms, 14 found no impact and 3 showed worsening of symptoms. Additionally, 12 studies only reported differences in hormone levels, and not the impact on AD symptoms. The symptoms measured varied between studies, reflecting disease-specific focus areas.

**Discussion:** This scoping review had broadly summarised the existing literature investigating the impact of oestrogen on three common ADs, suggesting that oestrogen may be anti-inflammatory within the context of ADs. Future research should investigate the combined influence of multiple hormones, across genders and assess their impact at repeated timepoints on ADs.

## Introduction

Symptoms of autoimmune disorders (ADs) vary both between disorders as well as between individuals. There is no known cure, and for many only limited treatments are available to manage symptoms. Between genders there are stark differences in AD experiences, with a much higher overall prevalence shown in women (∼80%), as well as differences shown in symptom severity and expression[1, 2]. Women have also reported changes in AD symptoms around reproductive events. For example, an increase in symptoms have been reported around menstruation, as well as a decrease in AD symptoms during pregnancy, with a marked increase immediately following birth[2-4]. Whilst the mechanisms behind such gender disparities are not fully understood, it has been suggested this may be due to differences in hormone levels, with varying hormone differences shown in women due to the biological potential for pregnancy[5]. The marked reduction of AD symptoms shown during pregnancy, has for example led to research investigating if administering reproductive hormones to individuals with multiple sclerosis could reduce symptoms[6, 7].

During pregnancy, significant biological and physiological changes occur to protect both the pregnant person and developing fetus, particularly in relation to hormones and the immune system[8]. These immune adaptations are essential to prevent the pregnant person’s immune system from overreacting and attacking the fetus[9]. Oestrogen, a sex hormone primarily responsible for regulating the female reproductive system, has been reported to exert anti-inflammatory effects[10, 11], and can therefore dampen the immune response. As ADs arise from an overactive immune system that attack healthy cells, the hormonal changes observed during pregnancy highlight a possible connection between other reproductive events and ADs. However, we currently do not understand if changes in AD symptom expression around reproductive events are related to individuals with ADs having different baseline hormone levels, making them more susceptible to hormone fluctuations, or if the hormone changes themselves directly cause symptom variation. Studies have examined the impact of administering varying levels of oestrogen (commonly available in hormone replacement therapies prescribed during menopause), on AD symptom expression[12-14]. Several studies have demonstrated oestrogen can exert both anti-inflammatory and neuroprotective effects for multiple sclerosis (MS)[3, 15]. However, research for women’s reproductive health, including the mechanisms contributing to poorer health outcomes, remains limited and underfunded, a consequence from a longstanding history of gender bias in medical research[16]. As a result, our understanding of these mechanisms, particularly those related to reproductive health, remain less evidenced.

Increased understanding within this area can clarify the mechanisms underlying variations in symptom expression across common ADs, highlighting contributing and modifiable pathways to disease symptomology. This study will systematically review the existing literature, to better characterise the relationship between oestrogen and common ADs. Greater insight into reproductive hormone mechanisms will help to enhance the knowledge of AD management around reproductive events, and importantly provide evidence to inform targeted care approaches.

## Method

This scoping review was conducted according to PRISMA (Preferred Reporting Items for Systematic Reviews and Meta-analyses) guidelines (Moher et al., 2009) and the protocol was preregistered on the Open Science Framework (https://osf.io/ge46a/). PubMed was searched until 23^rd^ November 2023 and a further search was conducted on 23^rd^ January 2025 to update the review with any relevant studies published following the initial search. Screening of eligibility was conducted by one reviewer (LS) at each stage, with a separate 20% check of articles by three additional reviewers completed at each stage: title and abstract, and full text. A further 100% data check was conducted by KEE. Any disagreements on eligibility were discussed and resolved by mutual consent.

### Inclusion criteria

The search strategy included key words (see supplementary methods) related to oestrogen, autoimmune disease, and specifically three top commonly reported autoimmune disorders (rheumatoid arthritis (RA), multiple sclerosis and type-one diabetes (T1D)); alternative ADs were not included to further refine the review. Oestrogen search terms included oestrogen, oestradiol and oestriol. During the initial screening stages, studies were excluded if they were review articles, or did not directly measure hormone amounts (Table 1). This was to refine the review away from less valid measurements of hormones, such as self-reported menarche for example which has been used as a proxy. Animal studies were also included. As the included ADs do not share a single primary symptom, specific symptom search terms were not included. This ensured broad data capture, reducing the risk of missing lesser known symptoms and associations. Instead, the strategy focused on identifying studies that measured oestrogen levels in participants with the specified ADs.

**Table 1:** Inclusion and exclusion criteria.

| Inclusion criteria | Exclusion criteria |
| --- | --- |
| Empirical research | Review articles |
| Peer reviewed publication | Conference abstracts, student theses |
| Oestrogen hormone | Only measured hormones other than oestrogens |
| AD measure of rheumatoid arthritis, multiple sclerosis or type one diabetes | Only measured ADs other than three in inclusion criteria |
| English language publication | Non-English publication |

### Data extraction

Data were extracted by one reviewer (LS) on study population location, design, sample size, age, AD measured, type of oestrogen measured, results, if animal study. To ensure accuracy, three independent reviewers independently reviewed 20% of studies at the title/abstract screening and full text inclusion stages. A 100% data check on data extraction was also completed by KEE, with an additional 20% of the dataset reviewed by two independent reviewers for accuracy.

## Results

The initial search (23/11/2023) identified 1162 studies after removal of duplicates (*n*=1). During the title/abstract screening 133 were included for full text screening. Of these, 55 were shown to be ineligible as they did not meet the inclusion criteria and were excluded (see Fig 1). Additionally, 12 articles were unavailable for full text, despite requesting a copy of the original text from the study author.

**Fig 1:**
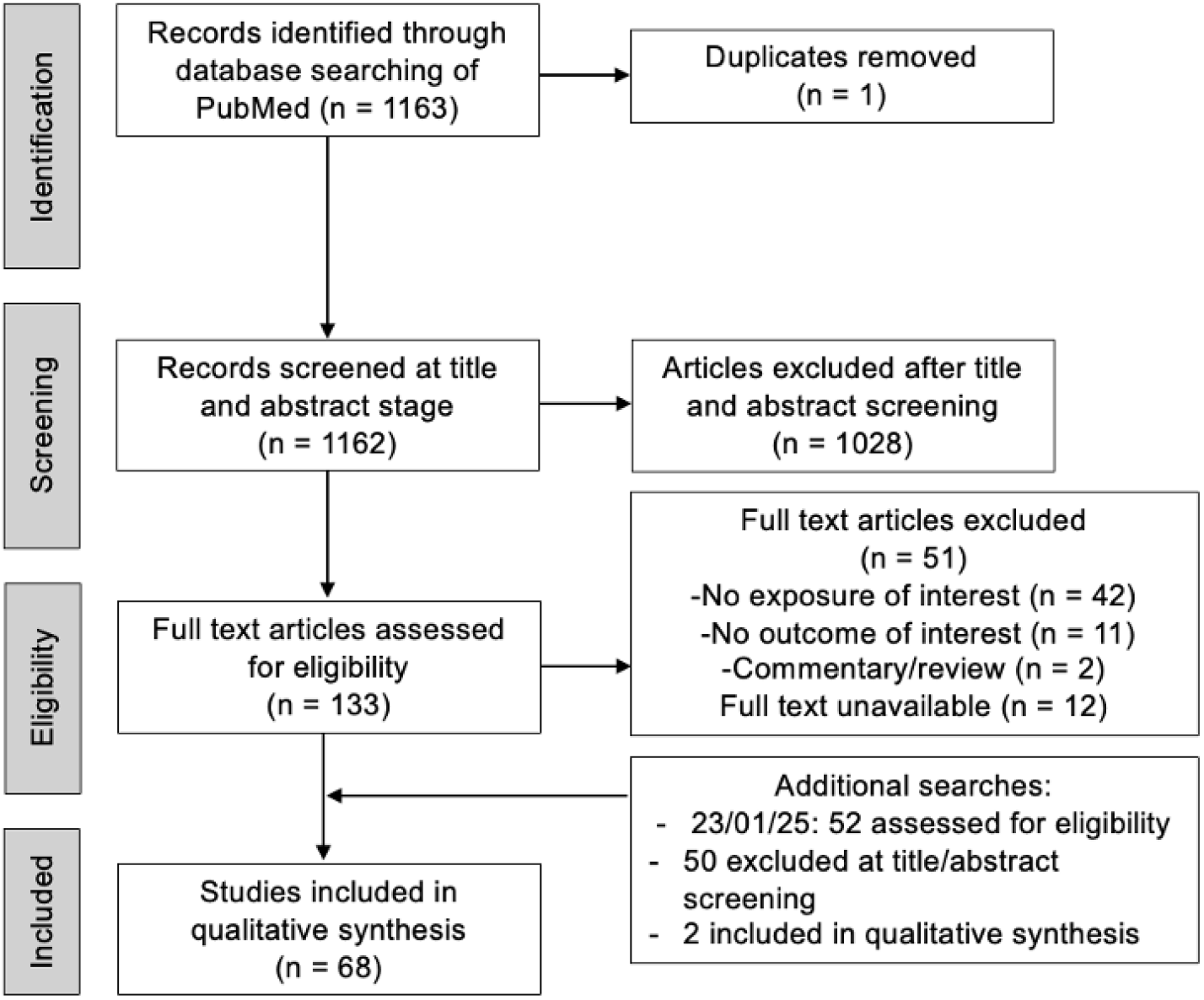
Flowchart of search strategy

An additional search was conducted on the 23/01/2025 to capture any articles published after the initial search. This found a further 52 studies, of which two were included after title/abstract and full text screening and subsequently included in the final review. The final searches resulted in 68 studies within the scoping review (see flowchart fig 1).

### Study characteristics

Of the final 68 included studies, 21 included populations from North America, 4 from South America, 6 from the UK, 27 Europe, 1 Australia, 1 Africa, 2 India, 1 Iran, 1 Korea and 1 Japan. Of the studies that reported sample sizes, these ranged from 10 to 381,306.

Of the included studies, 35 measured RA, 25 MS, and 7 type-one diabetes. Additionally, 1 study measured both MS and RA. Types of oestrogen hormones measured included oestrogen (8), oestradiol (55), and oestriol (5). Most studies investigated female participants only (50; 74%), 9 (13%) measured both genders and 8 (12%) measured males only. One animal study did not specify gender.

Within human studies, ages ranged from 14.9 (+/-2.3) to 70. In animal studies, rodents were aged 6-19 weeks. See Table 2 for percentages of animal and human studies.

**Table 2:** Demographics of included studies.

|  | Total (%) |
| --- | --- |
| **Sample gender** |  |
| Female only | 50 (74) |
| Male only | 8 (12) |
| Both | 9 (13) |
| NA | 1 (1) |
| **Sample animal/human** |  |
| Human | 42 (62) |
| Animal | 26 (38) |
| **Country** |  |
| Europe | 27 (40) |
| USA | 21 (31) |
| UK | 6 (9) |
| South America | 4 (6) |
| Iran | 3 (4) |
| India | 2 (3) |
| Korea | 1 (1) |
| China | 1 (1) |
| Australia | 1 (1) |
| Africa | 1 (1) |
| Japan | 1 (1) |

### Impact of hormones on AD

Of the 68 included studies, 12 did not measure the impact of oestrogen on AD outcomes and instead only reported the difference in hormone levels in those with ADs. Thirty four studies found that oestrogen hormones were associated with a reduction of or positive impact on AD symptom/incidence reduction, 14 showed no impact on AD symptoms and 3 gave a worsening of symptoms.

Of the studies that did not measure the impact of hormones on AD, and instead only presented hormone levels, 8 reported higher levels of hormones in those with ADs compared to controls, 5 lower levels and 4 showed no difference in levels. This higher level of hormones was mostly shown within RA. The direction of hormone levels varied within the studies including MS cases was dependent on which stage of the menstrual phase participants were in, with opposing effects shown for each phase (follicular or luteal) across the same number of studies (2). Four studies showed no difference.

Measured outcome types varied largely between studies (Fig 2) as expected due to the differences in commonly reported symptoms in the ADs measured. Within each AD there were themes of which outcome was measured. For example, in RA measures related to bone density, joint swelling/pain were mostly included. Within T1D, most studies measured hormone levels only (4/7) and the remaining measured glucose levels, incidence and bone mass. MS had the most varied outcome measures, including measures of incidence, disability scales, quality of life, brain lesions and fatigue (see Fig 2).

**Fig 2:**
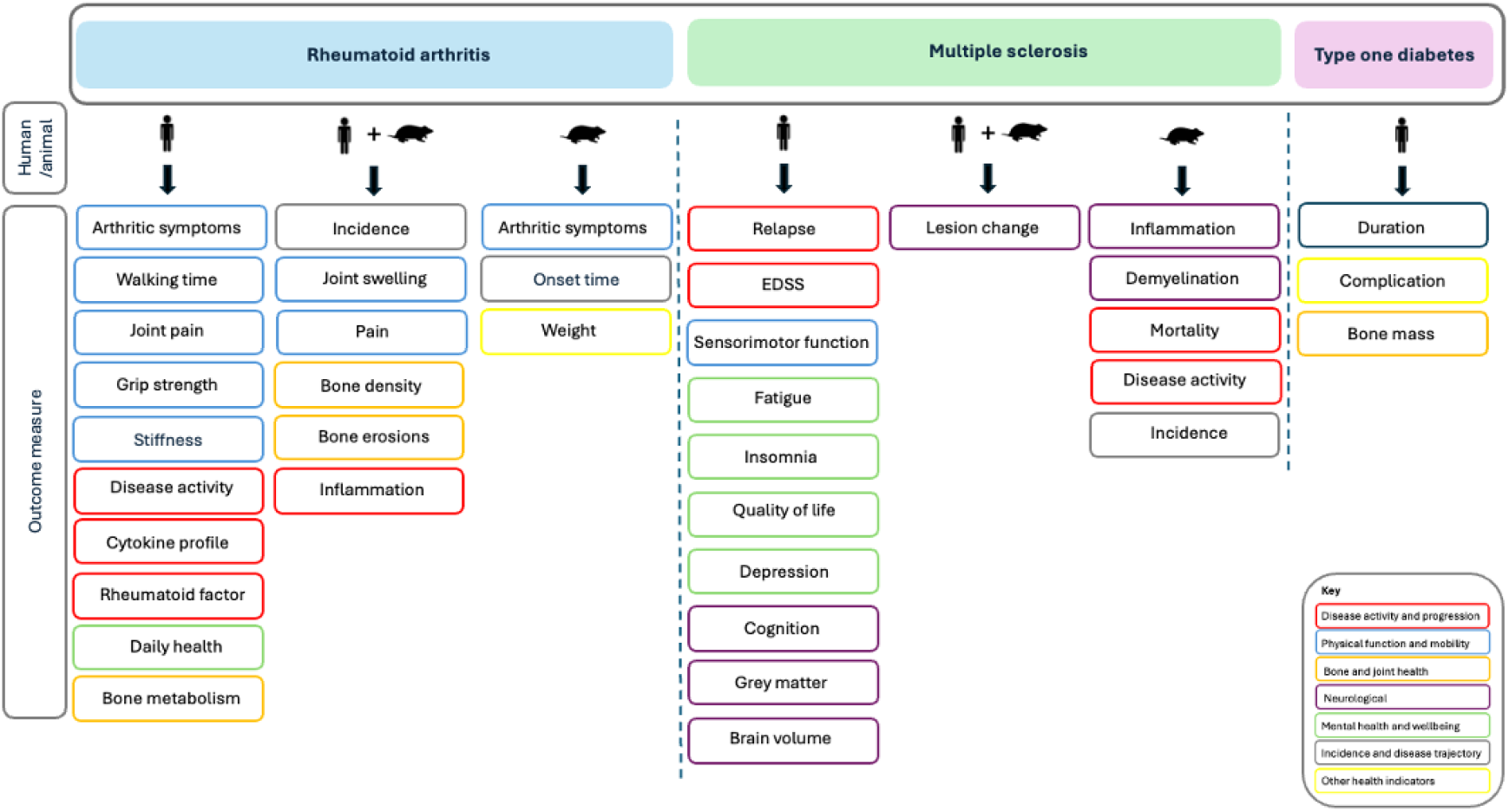
AD outcomes measured in studies

## Discussion

This scoping review has broadly summarised the published literature investigating the relationship between oestrogen hormones and three common ADs; RA, MS and T1D. The body of research evidenced in this review suggests that oestrogen may be anti-inflammatory within the context of ADs and is associated with a reduction in disease symptoms across multiple outcomes.

Interestingly, the studies that only measured hormone levels in individuals with an AD, rather than measuring hormone impact on symptoms, did not consistently show lower oestrogen levels compared to healthy controls. This suggests that lower oestrogen levels are not indicative of AD incidence. However, of the studies that only measured hormone amounts, it was shown that individuals living with an AD had a lower level of oestrogen hormones during a relapse compared to remission, which may be indicative of higher levels of oestrogen hormones being protective for individuals already living with an AD.

This review also strongly emphasises disparities between research focus within this area. For the measured ADs, RA was most often studied, and within oestrogen hormones, oestradiol was most often measured. The differences in type of measured ADs and hormones could be due to data availability or access, or due to limited scope of interest. Oestriol for example was included within the oestrogen search terms and only returned five studies included within the review. Oestriol is a hormone expressed at higher levels during pregnancy, and often less measured in non-pregnant samples, as reflected in the small amount of studies including this hormone. However, studies measuring the impact of administering this hormone to individuals living with MS has shown a vast reduction in symptoms and progression. Given the older age groups and focus on female participants for many included studies, it is also surprising that only four chose to specifically measure oestrone, a weak and readily available hormone for measurement. This hormone is often administered as part of hormone replacement therapy (HRT) treatment for menopause. Studies are therefore able to avoid ethical implications of administering hormones within human samples, and instead can collect information from women with an AD already receiving HRT within a menopausal sample. Future studies would benefit from the measurement of hormones commonly found in HRT.

Many included studies were conducted on animal models, where experimental manipulation of hormones and disease is less restricted. However, the type of hormones and symptom outcomes showed a similar pattern between human and animal studies. The limited scope may indicate a narrow overall research focus, potentially overlooking important interactions that could inform therapeutic advances.

A key strength of this scoping review is shown in the broad focus. By including research on multiple oestrogen hormones as well as three distinct ADs, this review captured a wide range of AD outcomes and their potential interaction with oestrogen. This approach reduced the risk of overlooking relevant findings that may have been missed if the search terms had been limited to specific AD symptoms. The importance of this broad search was particularly relevant for studies measuring MS, which showed the greatest amount of possible AD outcomes measured. Whilst MS shares commonly expressed symptoms for many, it is characterised by significant variability in symptoms experienced both across and within individuals[17]. Future research should aim to identify which specific aspect of AD activity are most impacted by hormones, using precise methodologies. Repeated hormone measurements across menstrual cycles for example, would also provide more valid and temporally sensitive data. Hormone levels are known to fluctuate across the life course and even within monthly cycles, therefore incorporating repeated measures would help control for these variations. Studies that did include data from multiple timepoints did show variation in symptom expression, however, limited studies included such longitudinal data.

Although not reported, some studies measured the impact of multiple hormones in addition to oestrogens. As hormone levels follow patterns of change alongside other hormones, for example across the menstrual cycle, future research should consider the combined impact of multiple hormones on AD symptoms. Future research investigating other reproductive hormones such as progesterone, or testosterone are welcomed to further discover how hormonal influences on AD disease symptomology and incidence is currently studied. Including testosterone for example, may have increased the number of studies investigating multiple genders, due to the heightened levels of this hormone expressed in males.

## Conclusion

This scoping review provides evidence for the interaction between oestrogen hormones and three commonly experienced ADs, and addresses a gap in the literature summarising the overall impact of oestrogen hormones on various outcomes of multiple ADs. The findings suggest a potentially anti-inflammatory effect of oestrogens on ADs. Importantly, it has shown the dominant areas where this research is focused, and highlight where future research is needed. For example, much of the research focused on oestradiol, as well as RA over other ADs. The majority of measured outcomes were for physical attributes, and less measured cognition and quality of life outcomes, which may overlook the internalising/wellbeing consequences which are also examples of disease progression in many ADs[18]. Further research would benefit from experimental studies investigating the impact of various reproductive hormones beyond oestrogen, across repeated timepoints.

## Data Availability

All data produced in the present work are contained in the manuscript.

## Acknowledgments and funding

The authors thank Evie Robins and Rafferty Welch from the University of Bristol, who supported the study in data quality checks and extraction.

KEE, is a member of the UK MRC Integrative Epidemiology Unit, this research was funded in part by the MRC (MC_UU_00032/7) and the University of Bristol. KEE is supported by funding from Bristol City Council, and the department for Education (G1058). For the purpose of open access, the author has applied a ‘Creative Commons Attribution (CC BY) public copyright licence to any Author Accepted Manuscript (AAM) version arising from this submission.

