## Supplementary material for "Oestrogen and autoimmune disease activity: A scoping review"

**Supplementary methods**

"autoimmune disorder OR “autoimmune disease” OR "multiple sclerosis" OR "rheumatoid arthritis OR type one diabetes OR type 1 diabetes

AND

*estrogen OR *estradiol OR *oestriol

**Supplementary Table 1**

| Citation | Population location | Autoimmune disease | Age | Animal/human sample | Sample size | Hormone | Gender | Outcome type | Descriptive results |
| --- | --- | --- | --- | --- | --- | --- | --- | --- | --- |
| [V Dziedziejko (2016)](https://academic.oup.com/endo/article/157/3/1013/2422581?login=false) | Sweden | RA | 8-10 weeks | Animal | na | Oestradiol | Female | Severity and frequency. Bone mineral density | Oestradiol alone and BZA/E2 combination reduced total arthritis severity over time (mean ± SEM): Vehicle 24.8 ± 3.5; E2 4.52 ± 2.0; BZA/E2 6.24 ± 2.6. |
| [Nielsen et al (2008)](https://pmc.ncbi.nlm.nih.gov/articles/PMC2384059/) | Germany | RA | na | Animal | 40 | Oestrogen | Female | Time-point of disease onset, incidence of manifest | Oestrogen delayed the average time-point of disease onset and reduced the number of diseased animals to 66% (p < 0.05 from day 13; NS at day 19). Oestrogen supplementation efficiently reduced paw inflammation, decreasing paw volume by 48% on day 28 (p < 0.01) |
| [Kim et al (1999)](https://www.neurology.org/doi/10.1212/wnl.52.6.1230?url_ver=Z39.88-2003&rfr_id=ori:rid:crossref.org&rfr_dat=cr_pub%20%200pubmed) | USA | MS | 6-8 weeks | Animal | 106 | Oestriol | Both | Incidence and lesions | Estriol ameliorated EAE symptoms. Pellets at 5, 15, and 30 mg significantly reduced clinical severity (p < 0.0005, p < 0.005). Post onset oestriol treatment after disease onset also significantly lessened severity compared to placebo (p < 0.05) |
| [Garay et al (2010)](https://pubmed.ncbi.nlm.nih.gov/25961971/) | South America | MS | 9-11 weeks | Animal | na | Oestradiol | Female | White matter demyelination | Maximum clinical scores were 2.9-fold higher in EAE mice compared to EAE + estradiol + progesterone mice (p < 0.05). Co-treatment with E2 and progesterone dramatically reduced EAE demyelination (p < 0.001, p < 0.01) |
| [Gourdy et al (2016)](https://academic.oup.com/endo/article/157/1/258/2251847?login=true) | California | T1D | na | Animal | na | Oestradiol | Female | Incidence | Oestradiol showed protective effects at weeks 4–12 (p < 0.02) and 4–20 (p < 0.001). Ovariectomised mice had significantly delayed diabetes onset at 16–18 weeks compared to sham (p < 0.04), but this effect did not persist at 30 weeks. Sham vs. oestradiol was significant at 12–28 weeks (p < 0.015) and between 12–20 vs. 12–28 weeks (p < 0.003), but not significant between sham and oestradiol at 12–20 weeks (NS) |
| [Meyer et al (2023)](https://www.sciencedirect.com/science/article/pii/S0023683723001320?via%3Dihub) | USA | MS | 8-16 weeks | Animal | 36 | Oestriol | Female | Demyelination, brain atrophy | Oestriol-treated EAE mice showed significantly reduced cerebral atrophy compared to placebo-treated EAE mice (p = 0.012). Cortical layer V neurons were preserved in oestriol-treated mice versus placebo (p = 0.0047), with normal spine density maintained (p = 0.044). Microglial activation was also reduced in the cerebral cortex of treated mice compared to placebo (p = 0.0062) |
| [Stubelius et al (2011)](https://www.sciencedirect.com/science/article/pii/S1521661611000763?via%3Dihub) | Denmark | RA | na | Animal | 37 | Oestradiol | Female | Paw swelling | Oestradiol significantly decreased the incidence of arthritis compared to placebo-treated mice (p < 0.01) and reduced severity scores (p < 0.05) |
| [Ganesan (2008)](https://www.sciencedirect.com/science/article/pii/S8756328208002810?via%3Dihub) | Chennai | RA | na | Animal | 66 | Oestradiol | Female | Paw swelling, volume and edema | In ovariectomised (OvX) female RA rats, oedema was significantly suppressed by oestrogen (p < 0.01) |
| [Subramian et al (2005)](https://www.sciencedirect.com/science/article/pii/S1521661605000306?via%3Dihub) | USA | RA | na | Animal | 7 | Oestradiol | Male | Arthritic symptoms | 200ug of EE signifcantly reduced joint inflamation (p<.05) |
| [Jahantigh et al (2023)](https://pmc.ncbi.nlm.nih.gov/articles/PMC10008397/) | Iran | RA | 6 weeks | Animal |  | Oestradiol | Male | Paw swelling and stiffness | The average mean arthritis index significantly regressed in RA rats receiving oestrogen treatment (p < 0.05) |
| [Subramanian et al (2003)](https://journals.aai.org/jimmunol/article/170/3/1548/71207/Oral-Feeding-with-Ethinyl-Estradiol-Suppresses-and) | Maine | MS | 6-7 weeks | Animal | 42 | Oestradiol | Female | Peak clinical disease score, cumulative disease index, incidence and mortality rates, demylination and inflamation of spinal cord | Mice fed oestrogen had delayed disease onset (19.5 ± 0.7 days) compared to controls (13.3 ± 1.6 days) and exhibited less severe disease, with a lower peak score (0.6 ± 1.4 vs. 3.0 ± 1.2; p < 0.05 for both). Oestrogen treatment also reduced disease incidence (4/14 vs. 16/16) and cumulative disease index (3.9 ± 9.6 vs. 27.5 ± 10.7). Histopathological analysis showed reduced inflammation and demyelination in the spinal cord |
| [Saravia et al (2004)](https://onlinelibrary.wiley.com/doi/full/10.1111/j.1365-2826.2004.01223.x?saml_referrer=) | Argentina | T1D | na | Animal | 14 | Oestradiol | Male | Blood glucose levels | Blood glucose levels measured 20 days after diabetes induction were slightly lower in oestrogen-treated mice (601 ± 24.4 mg/dl) compared to untreated mice (654 ± 76.4 mg/dl) (p > 0.05) |
| [Santora et al (2007)](https://pubmed.ncbi.nlm.nih.gov/17526806/) | USA | RA | na | Animal | 32 | Oestradiol | Female | Paw volume, weight | Hormone treatments did not influence paw volume by day 7. However, by day 14, all treatments significantly decreased primary paw inflammation compared to vehicle controls (p ≤ 0.05), and this reduction persisted through day 21 (p ≤ 0.05). Treatment with oestradiol alone further reduced body weight compared to controls (p < 0.05) |
| [Jochems et al (2007)](https://onlinelibrary.wiley.com/doi/10.1002/art.22873) | Denmark | RA | na | Animal | 90 | Oestradiol | Female | Bone mineral density | Oestradiol administered prophylactically delayed arthritis onset (p < 0.05), though the effect was not sustained. Oestradiol significantly reduced disease severity (p < 0.001) and protected against paw and cartilage destruction (p < 0.001 for both), but had no significant effect on bone resorption or formation (significance not reported) |
| [Mattson et al (1991)](https://pmc.ncbi.nlm.nih.gov/articles/PMC1535724/) | UK | RA | na | Animal | 17 | Oestradiol | Female | Paw swelling | Oestradiol treatment compared to control showed a median change in clinical scores of 1.46 ± 0.09 (p < 0.01 |
| [Santora et al (2005)](https://pubmed.ncbi.nlm.nih.gov/15956749/) | USA | RA | na | Animal | 32 | Oestradiol | Female | Paw swelling, bone and soft tissue changes | Weekly treatment with oestradiol valerate decreased tissue damage in the injected hind paw (mean ± SEM: 11.2 ± 0.7) compared to vehicle-treated rats (16.3 ± 0.8; p < 0.05). Combined treatment with oestradiol valerate and relaxin was more effective than either treatment alone (p < 0.05) |
| [Haghmorad et al (2016)](https://www.tandfonline.com/doi/full/10.1080/1547691X.2016.1223768) | Iran | MS | 10 weeks | Animal | ~50 | Oestradiol | Female | Disease accumulation of movement problems, incidence, day of onset, day of maximal clinical signs, and total disease score. Demyelination of the spinal cord | Control group (placebo): 100% incidence; onset 10.6 ± 0.5 days; max score 4.7 ± 0.5; mean score 4.1 ± 0.3; CDI 38.8 ± 1.5. Low treatment (0.25 mg): 100% incidence; onset 10.6 ± 0.6 days; max score 4.2 ± 0.3; mean score 3.6 ± 0.4; CDI 33.5 ± 1.45. Medium treatment (2.5 mg): 70% incidence (p < 0.05); onset 12.1 ± 0.4 days; max score 1.5 ± 0.5 (p < 0.001); mean score 0.9 ± 0.3 (p < 0.001); CDI 12.2 ± 0.64 (p < 0.001). High treatment (15 mg): 65% incidence (p < 0.05); onset 12.6 ± 0.6 days; max score 1.4 ± 0.5 (p < 0.001); mean score 0.7 ± 0.2 (p < 0.001); CDI 10.1 ± 0.55 (p < 0.001). Pregnancy-level oestradiol significantly reduced incidence (p < 0.05) and severity (p < 0.001); medium levels reduced clinical signs (p < 0.05) |
| [Inoue et al (2013)](https://www.sciencedirect.com/science/article/pii/S0006291X13005901?via%3Dihub) | Japan | RA | 8 weeks | Animal | 12 | Oestradiol | Female | Incidence and bone destruction/erosion | In sham-operated mice, both the fifth metatarsal bone and calcaneus were destroyed, whereas in oestradiol-treated mice, only the fifth metatarsal bone was damaged. In sham-operated mice, both the fifth metatarsal bone and calcaneus were destroyed, whereas in oestradiol-treated mice, only the fifth metatarsal bone was damaged. |
| [Palanszynski et al (2004)](https://www.sciencedirect.com/science/article/pii/S016557280300571X?via%3Dihub) | USA | MS | 6-8 weeks | Animal | N/A | Oestriol | Both | Disease severity, average peak score, cumulative disease index | Disease severity was significantly reduced in females treated with estriol compared to placebo (p < 0.0001); a significant reduction in disease severity was also observed in males treated with estriol (p < 0.0001); both mean peak disease scores and cumulative disease index were significantly decreased by estriol treatment. |
| [Jansson et al (1994)](https://www.sciencedirect.com/science/article/pii/0165572894900302?via%3Dihub) | Germany | RA +MS | 8-12 weeks | Animal | 77 | Oestradiol and oestriol | Female | Clinical severity | Oestradiol treatment delayed disease onset by 35 days (p < 0.005), but disease developed with similar frequency and severity to controls in naïve rats. A low dose of oestradiol was less effective than a high dose, delaying onset by 10 days (p < 0.01). Estriol was as effective as high-dose oestradiol, delaying onset by 25 days compared to controls (p < 0.01). Oestradiol pellets completely blocked CIA during treatment, but disease developed after treatment ended. |
| [Jansson et al (1992)](https://pmc.ncbi.nlm.nih.gov/articles/PMC1554486/) | Sweden | RA | 6-12 weeks | Animal | 127 | Oestradiol | Both | Joint swelling | Male mice required less CII to develop CIA, showed earlier onset, and reached higher frequency and severity compared to females. Oestradiol suppressed arthritis frequency in both castrated (p < 0.05) and normal (p < 0.05) female mice compared to controls. A 3.2 µg dose suppressed both frequency and severity of arthritis in castrated (p < 0.05) and normal (p < 0.05) male mice. A 0.2 µg dose did not affect frequency but significantly reduced severity in both normal (p < 0.05) and castrated (p < 0.05) male mice. |
| [Jochems et al (2010)](https://pmc.ncbi.nlm.nih.gov/articles/PMC3009959/) | Sweden | RA | 7-19 weeks | Animal | 44-60 | Oestradiol | Female | Joint swelling | Oestradiol treatment resulted in 20% arthritis onset and a slower disease progression compared to vehicle controls (p < 0.001). Joint destruction was significantly lower in oestradiol-treated mice (p < 0.01) and sham-operated mice (p < 0.01). Endogenous oestrogens showed a protective effect against bone mineral density (BMD) loss (p < 0.05, p < 0.001), with oestradiol-treated mice displaying the highest BMD (p < 0.001). |
| [Engdahl et al (2010)](https://onlinelibrary.wiley.com/doi/abs/10.1002/art.25055) | Denmark | RA | 3 months | Animal | 78-90 | Oestradiol | Female | Joint swelling | Oestradiol delayed the onset of arthritis and reduced disease severity, decreased arthritis frequency from day 27 (p < 0.05), and led to significant amelioration of arthritis from day 32 (p < 0.01) compared to controls. |
| [Subramanian et al (2005)](https://www.sciencedirect.com/science/article/pii/S1521661605000306?via%3Dihub) | USA | RA | NA | Animal | 40 | Oestradiol | Male | Joint swelling | A 50 µg dose of ethinylestradiol effectively suppressed arthritic symptoms within 10 days of treatment initiation, while a 200 µg dose had a stronger effect, producing significant daily improvements within 3 days of treatment (p < 0.05). |
| [Ganesan et al (2008)](https://www.sciencedirect.com/science/article/pii/S8756328208002810?via%3Dihu) | India | RA | NA | Animal | 66 | Oestradiol | Both | Paw volume, radiology, histopathology of joint, markers for bone turnover | Oestrogen admininstration suppresed severe edema of ovx rats (p < 01) |
| [Hoffman et al (2001)](https://www.sciencedirect.com/science/article/pii/S0014488601977830?via%3Dihub) | USA | MS | NA | Animal | 25 | Oestradiol | Female | Sensorimotor function and inflammation | Oestrogen replacement in OVX MBP animals largely improved sensorimotor function, with 5 out of 7 animals showing no or minimal signs of EAE. Estrogen alone reduced spinal cord inflammation (no p value reported) |
| [Mackenzie-Graham et al (2018)](https://pmc.ncbi.nlm.nih.gov/articles/PMC6160650/#brb31086-sec-0006title) | USA | MS | 20-53. Mean 37.3 years | Human | 111 | Oestriol | Female | Grey matter in brain and cognitive scores | The estriol plus glatiramer acetate (GA) group showed significantly less annualized whole grey matter loss (0.5%) compared to the placebo plus GA group (1.5%) (p = 0.04). Improvement in PASAT3 performance was observed in the estriol plus GA group versus placebo plus GA (p = 0.02), with a trend toward improvement on PASAT2 (p = 0.07). No baseline differences were found in gadolinium-enhancing lesions, FLAIR lesion volume, whole brain volume, white matter volume, cortical grey matter volume, or voxelwise grey matter volume |
| [Martinez et al (2016)](https://onlinelibrary.wiley.com/doi/abs/10.1111/dme.13078) | Chile | T1D | Mean 13.9 years | Human | 70 | Oestradiol and oestrone | Female | Hormone levels | Adolescents with Type 1 diabetes had significantly lower serum oestradiol (E2) and free E2 levels during the follicular phase compared to controls (p < 0.01). Additionally, E1 levels were significantly higher in adolescents with Type 1 diabetes during the luteal phase (p < 0.01). Regression analysis showed a negative association between oestradiol and Type 1 diabetes (β = −24.8, SE = 7.5, p = 0.002). |
| [Trenova et al (2013)](https://www.tandfonline.com/doi/full/10.1179/1743132812Y.0000000120) | Bulgaria | MS | 19-49 years | Human | 35 | Oestradiol | Female | Relapse and remission. EDSS | In patients with low hormone levels, estradiol increased during remission compared to relapse (mean ± SE = 207.90 ± 39.70; SD = 177.50; t = 9.45, p < 0.01). In patients with normal hormone levels, no difference was observed between remission and relapse (t = 0.50, p > 0.05). No differences among groups were found in disability as measured by EDSS |
| [Talaat et al (2018)](https://pmc.ncbi.nlm.nih.gov/articles/PMC6223740/) | Egypt | MS | 16-35 years | Human | 40 | Oestrogen | Female | EDSS | Oestrogen levels were 1.84 ± 0.27 pg/ml (mean ± SD) compared to controls at 1.74 ± 0.2 pg/ml (p = 0.12). The correlation between oestrogen levels and EDSS was r = −0.007, p = 0.96. |
| [Vukusic et al (2020)](https://journals.sagepub.com/doi/10.1177/1352458520978218?url_ver=Z39.88-2003&rfr_id=ori:rid:crossref.org&rfr_dat=cr_pub%20%200pubmed) | France Italy | MS | Mean 32.1. 23.9-42.3 years | Human | 200 | Oestradiol | Female | Postpartum MS activity of annualized relapse rate | Annualized relapse rate showed NS difference between steroid and placebo groups from 0–12 weeks postpartum (p = 0.79), 12–24 weeks (p = 0.73), and 0–24 weeks (p = 0.95). |
| [Hassan et al (2013)](https://pmc.ncbi.nlm.nih.gov/articles/PMC5124523/) | USA | T1D | 18-50 years | Human | 165 | Oestradiol | Female | Hormone levels only | No difference in mean total or bioavailable estradiol between individuals with and without Type 1 diabetes in unadjusted or adjusted models (p > 0.74). Total estriol (pg/ml) means (SE): diabetes 301.5 (137.1), controls 294.3 (117.7); unadjusted difference 6.0 (p = 0.74), adjusted difference 4.8 (p = 0.79) |
| [Koepsell et al (1994)](https://academic.oup.com/ije/article/23/6/1248/660392) | USA | RA | 25-64 years | Human | 727 | Oestrogen | Female | RA initiation | Age-adjusted relative risk (RR) for women ever using non-contraceptive oestrogens: 1.04 (95% CI: 0.70–1.55) |
| [Bijlsma et al (1987)](https://pmc.ncbi.nlm.nih.gov/articles/PMC1003387/?page=1) | USA | RA | na | Human | 10 | Oestradiol | Female | Walking time, joint tenderness, grip strength | Joint tenderness (mean ± SE) was 26.9 ± 13.2 for oestrogen and 0.6 ± 5.8 for placebo (p = 0.1). Swollen joints were −1.5 ± 7.8 for oestrogen and 11.2 ± 9.9 for placebo (p > 0.5). Grip strength increased by 15.0 ± 5.7 in the oestrogen group and 19.9 ± 10.05 in placebo (p = 0.11). 30m walking time improved by −9.7 ± 4.2 seconds with oestrogen and worsened by 3.4 ± 2.5 seconds with placebo (p = 0.03) |
| [Zakrzewska-Pniewska et al (2011)](https://www.sciencedirect.com/science/article/abs/pii/S0028384314601201) | Poland | MS | 19-65 years | Human | 96 overall. 46 with MS | Oestradiol | Female | Hormone levels, EDSS | Mean oestrogen levels were 137.7 ± 144.26 pg/mL for oestradiol. In the follicular phase, controls had 102 pg/mL and MS patients 179.13 pg/mL. In the luteal phase, controls had 122 pg/mL and MS patients 231.29 pg/mL. Oestradiol levels were abnormal in 12 patients. |
| [Wei and Lightman (1997)](https://academic.oup.com/brain/article/120/6/1067/300553?login=true) | London | MS | 24-70 years | Human | 26 | Oestradiol | Female | Hormone levels | 4/16 (25%) premenopausal MS patients had low oestradiol; 6 postmenopausal patients not on HRT all had low oestradiol consistent with menopausal status; no significant hormone differences between patients with and without fatigue (data not shown) |
| [Tengstrand et al (2003)](https://www.jrheum.org/content/30/11/2338.long) | Sweden | RA | 23-70 years | Human | 230 | Oestradiol and estrone | Male | Hormone levels, inflammatory activity, disease activity score in 28 joints | Oestrodiol concentrations with RA means and interquartile ranges (93(77-128) p<0.0001, controls 73.5(56-95). Correlations with symptoms: oestradiol and DAS (disease activity) p < 0.0001, R = 0.38. patients global assessment VAS of disease activity p<0.5; swollen joint count p<0.001; tender joint count p=0.08; ESR (inflamation) p<0.001, r=0.49, ; HAW p<0.05, r=0.25. |
| [van den Brink et al (1993)](https://pmc.ncbi.nlm.nih.gov/articles/PMC1005630/) | netherlands | RA | 61-83 years | Human | 40 | Oestradiol | Female | Bone metabolism and bone mineral density | Oestrogen group showed decreased osteoblastic activity (p < 0.005); lumbar spine BMD increased from 0.96 (0.17) at start to 1.00 (0.07) after 6 months vs. placebo (p < 0.005); left femoral neck BMD in oestrogen group changed from 0.63 (0.08) to 0.65 (0.08) (p < 0.05) compared to placebo starting at 0.70 (0.11) and 0.69 (0.11) at 6 months; femoral neck in placebo group changed from 0.69 (0.10) to 0.67 (0.11) |
| [Tomassini et al (2005)](https://pmc.ncbi.nlm.nih.gov/articles/PMC1739476/) | Italy | MS | mean 32.3 | Human | 96 | Oestradiol | Both | EDSS, MRI lesion changes | Oestradiol increased significantly during remission vs relapse in patients with low hormone levels (mean ± SE 207.90 ± 39.70; SD 177.50, t=9.45, p<0.01). For normal hormone levels, no difference in remission vs relapse was found |
| [Hall et al (1993)](https://pubmed.ncbi.nlm.nih.gov/8484674/) | UK | RA | 45-65. 56 mean | Human | 703 | Oestradiol | Female | Bone mineral density. Erythrocyte sedimentation rate (inflammation) | Oestradiol levels were 81.4 (150.5) in never-steroid users and 52.8 (105.8) in controls, p = NS. Correlations of oestradiol with bone mineral density were 0.28 (spine, p < 0.01), 0.29 (femur, p < 0.01), and 0.42 (whole body, p < 0.01). No significant correlations were found with early morning stiffness (-0.16, p > 0.05), erythrocyte sedimentation rate (-0.17, p > 0.05), or VAS pain (-0.11, p > 0.05) |
| [Hylmarova et al (2020)](https://www.jstage.jst.go.jp/article/endocrj/67/1/67_EJ19-0280/_html/-char/en) | Czech Republic | T1D | 18-50 (mean 33) | Human | 57 | Oestradiol | Male | Duration of disease, diabetic complication | Control men (n=150) had median estradiol levels of 90.9 pg/mL (reference range 41.4–159.0). Diabetic men (n=41) showed 8 (19.5%) with low estradiol, 1 (2.4%) with high estradiol, median 67.5 pg/mL, IQR 51.0; test statistic = –1.899, p = 0.058 compared to controls. No association was found between estradiol concentration and duration of diabetes |
| [Rakic et al (2006)](https://link.springer.com/article/10.1007/s00125-006-0154-2) | Australia | T1D | male and female means rangind between 43.4 to 67.2 | Human | 70 | Oestradiol | Both | Hormone levels | Females: Type 1 diabetes 212 pmol/L (range 52–859), Type 2 diabetes 18 pmol/L (range 6–54), p < 0.001. Maleas: Type 1 diabetes 86 pmol/L (range 49–151), Type 2 diabetes 61 pmol/L (range 23–163), p = 0.15 |
| [Aroso Dias et al (1989)](https://link.springer.com/article/10.1007/BF02207241) | Portugal | RA | means 47.2-50.9 | Human | 36 | Oestradiol | Female | Hormone levels | Serum oestradiol levels in RA cases: mean 37.6; controls: mean 42.8; NS |
| [Salliot et al (2022)](https://www.sciencedirect.com/science/article/pii/S1297319X22000331?via%3Dihub) | France | RA | 40-65 (mean 49) | Human | 637 cases. 78391 total | Oestrogen | Female | Prevalance | High oestrogen exposure was inversely associated with risk of rheumatoid arthritis in postmenopausal women compared to low exposure, with a multivariable hazard ratio of 0.37 (95% CI 0.2–0.8, p < 0.05) versus low exposure |
| [Soto et al (2011)](https://www.sciencedirect.com/science/article/pii/S1056872709001172?via%3Dihub) | Chile | T1D | 14-32 | Human | 95 | Oestradiol | Female | Bone mass | Oestradiol means (SD): Adolescents with diabetes 68.3 ± 34 pg/ml, controls 51.4 ± 17 pg/ml; adults with diabetes 69.3 ± 34 pg/ml, controls 74.9 ± 27 pg/ml; all p > 0.05 |
| [Hall et al (1994)](https://pmc.ncbi.nlm.nih.gov/articles/PMC1005262/) | UK | RA | 46-65 | Human | 168 | Oestradiol | Female | Erythrocyte sedimentation rate (ESR), articular index (AI), visual analogue pain scale (VPS) and early morning stiffness | Placebo-treated patients had higher early morning stiffness scores (p = 0.02). No overall difference in disease activity changes. Only 58% of HRT patients had estradiol >100 pmol/l at 3 or 6 months. Compliers showed improved ESR at 3 months (p = 0.04), lower VPS vs placebo (p < 0.05), and improved arthritis index (p < 0.01) placebo reported higher EMS scroe (*p*=.02); no signif diff in changes of idsease activity parameter overall; oly 58% if the HRT group had E2 levels greater than 100pmol/l at3 or 6 months; the compliers had signif improvements in ESR at 3 months (*p*=.04) adn VPS comapred to placebo (*p*<.05) adn AI (*p*<.01). |
| [Voshkuhl et al (2016)](https://www.sciencedirect.com/science/article/pii/S1474442215003221?via%3Dihub) | USA | MS | 18-50 | Human | 158 | Oestriol | Female | Relapse rate | After 24 months, relapse rate was 0.25/year in estriol vs. 0.37/year placebo (p=0.077). Annualized relapse rate was lower at 12 months (0.25 vs. 0.48, p=0.016) but not at 24 months (p=0.098). Fatigue improved with estriol at 24 months (p=0.009). Cognition was better at 12 months (p=0.054) but not 24 months (p=0.902) |
| [Foroughipour wt al (2012)](https://pmc.ncbi.nlm.nih.gov/articles/PMC3697216/) | Iran | MS | 15-35 | Human | 46 | Oestradiol | Female | EDSS score | Oestradiol level was only lower in follicular phase in patients vs controls (p = 0.026). Inverse correlation between EDSS score and oestradiol (r = –0.471, p = 0.008) |
| [van den Brink et al (1993)](https://pmc.ncbi.nlm.nih.gov/articles/PMC1005216/) | The Netherlands | RA | Mean 61-63 | Human | 40 | Oestradiolvalerate | Female | Joint inflammation, pain, erythrocyte sedimentation rate | No difference in any disease parameter between groups at 6 months (p = 0.23) or 12 months (p = 0.07) |
| [Bansil et al (2009)](https://onlinelibrary.wiley.com/doi/abs/10.1111/j.1600-0404.1999.tb00663.x?sid=nlm%3Apubmed) | USA | MS | Mean 36.9-42.5 | Human | 30 | Oestradiol | Female | MRI lesions | Higher oestradiol levels correlated with greater number of gadolinium-enhancing lesions (p = 0.04) |
| [Castagnetta et al (2003)](https://www.jrheum.org/content/30/12/2597.long) | caucasian Italian | RA | 40-66 (mean 54.5) | Human | 12 | Oestrone | Both | Hormone levels only | Synovial fluid oestrogen levels slightly elevated in RA vs controls (NS); free estrone levels higher in RA vs controls (p = 0.035) |
| [Yuk et al (2023)](https://www.sciencedirect.com/science/article/pii/S0049017223001221?via%3Dihub) | Korea | RA | mean 50 (47-54) | Human | 277,982 | Oestradiol | Female | Incidence | NS association between menopausal hormone therapy (MHT) and RA risk (p = 0.054); oestrogen alone also not significant (p = 0.556). MHT use <3 years associated with increased RA risk (p < 0.001), but oestrogen alone not associated (p = 0.199) |
| [Cutolo et al (1988)](https://onlinelibrary.wiley.com/doi/epdf/10.1002/art.1780311015) | Italy | RA | Mean 60 | Human | 22 | Oestradiol | Male | Hormone levels only | Basal oestradiol concentrations not different from controls, p value not reported |
| [Doran et al (2003)](https://www.jrheum.org/content/jrheum/31/2/207.full.pdf) | USA | RA | Mean 57.5 | Human | 890 | Oestrogen | Female | Incidence | Ever use of oral contraceptives associated with reduced RA risk (p = 0.02); similar association in women with ≥6 months use (p = 0.02); no association with current use (p = 1.00); protective effect remained after adjustment for age and smoking |
| [Triantafyllou et al (2015)](https://www.tandfonline.com/doi/full/10.3109/00207454.2015.1069825) | Greece | MS | Mean 39.06 | Human | 133 | Oestradiol | Both | EDSS | Mean total oestradiol levels in postmenopausal women inversely associated with disability; lower E2 linked to greater neurological impairment but not significant (p = 0.095) |
| [MacNonald et al (1994)](https://pmc.ncbi.nlm.nih.gov/articles/PMC1005244/) | UK | RA | 53-55 | Human | 62 | Oestradiol | Female | Bone mineral density | In the HRT group, subjective overall well-being improved at weeks 12 and 48 (p < .05). No lab differences except increased C-reactive protein (p < .05). Oestradiol increased significantly (p < .01). Lumbar spine BMD increased significantly (p < .03); no significant change in femoral neck or wrist BMD |
| [De Giglio et al (2016)](https://link.springer.com/article/10.1007/s40263-016-0401-0) | Italy | MS | 18-45 | Human | 149 | Oestradiol | Female | Cognitive impairment. Fatigue, MS quality of life, depression | At 12 months, no difference in cognitive impairment between groups (p = .24). At 24 months, cognitive impairment was lower in the 40 mcg E2 group compared to control (p = .03). Mood and fatigue scores were comparable across the groups over time at both time points. However, at month 24, group 3 showed worsening on the sexual function subscale of the MS quality-of-life questionnaire (p = 0.03) |
| [Walitt et al (2009)](https://pmc.ncbi.nlm.nih.gov/articles/PMC2661110/) | USA | RA | Mean 64.1. 50-79 | Human | 27347 | Oestrogen | Female | RA incidence and severity of joint pain | NS difference in RA incidence between PHT and placebo (p=.134); adjusted reduction NS (p=.065). RA patients on PHT improved 7% vs 5.5% deterioration on placebo (p=.04), lost after adjustment (p=.33). Estrogen alone showed persistent protective effect on PCS (p=.009, p=.055) |
| [Tengstrand and Hafstrom (2002)](https://www.jrheum.org/content/jrheum/29/11/2299.full.pdf) | Sweden | RA | Mean 57.5 | Human | 104 | Oestradiol, oestrone | Male | Bone mineral density, bone erosions. | No difference in oestradiol and oestrone levels or BMD at all sites (no p value provide) |
| [Rovensky et al (2005)](https://pubmed.ncbi.nlm.nih.gov/15971415/) | Slovakia | RA | Males: mean 52.4. Women: 47.8 | Human | 50 | Oestradiol | Both | Hormone levels only | Oestradiol levels were higher in RA than OA patients (females p<.05, males p<.01). Synovial oestradiol correlated with synovial fluid cell count (inflammation) (p<.021) |
| [Cevik et al (2004)](https://onlinelibrary.wiley.com/doi/full/10.1111/j.1368-5031.2004.00005.x?saml_referrer) | Turkey | RA | 22-70. ean 45.68 | Human | 82 | Oestradiol | Female | Hormone levels only | NS difference in oestradiol concentration between RA patients and controls across all menopause stages (p > .05) |
| [Kim et al (2021)](https://pmc.ncbi.nlm.nih.gov/articles/PMC8118114/) | USA | MS | Mean 67.5 | Human | 381306 | Oestradiol | Female | Incidence | Hormone therapy use lowered MS risk (p<.001), with oestradiol formulations showing greater reduction (p<.001) |
| [Hall et al (1994)](https://onlinelibrary.wiley.com/doi/abs/10.1002/art.1780371014) | UK | RA | 45-65. Mean 55.8 | Human | 218 | Oestradiol | Female | Bone mineral density | Mean change in spinal BMD was significantly greater in HRT vs calcium-treated patients at 12 months (p=.01) and 24 months (p<.001), with HRT increasing BMD. No difference in femoral BMD at either time point (p not provided) |
| [Bove et al (2017)](https://journals.sagepub.com/doi/10.1177/1352458517692420?url_ver=Z39.88-2003&rfr_id=ori:rid:crossref.org&rfr_dat=cr_pub%20%200pubmed) | USA | MS | Mean 31.4-40.3 (across three groups) | Human | 162 | Oestrogen OC | Female | Relapse rate EDSS | Annualized relapse rate showed no difference across OC groups (p=.057) but was lower in past versus never users (p=.031). Over 8 years, never-users had a non-significant greater increase in EDSS compared to current and past users (p=.28) |
| [Juutinen et al (2022)](https://www.msard-journal.com/article/S2211-0348(22)00606-X/fulltext) | Finland | MS | Mean 52.1. 48-54 | Human | 27 | Oestradiol | Female | Vasomotor frequency, depressive symptoms insomnia, cognitive performance | No change in EDSS and SDMT scores after 12 months (p = .56 and p = .33). Depression, insomnia, and auditory processing scores decreased (p = .017, p = .05, p = .0002), but SDMT showed no change (p = .33) |
| [Bove et al (2016)](https://pmc.ncbi.nlm.nih.gov/articles/PMC5075979/#s1title) | USA | MS | Mean 56 | Human | 95 | Oestrogen | Female | Physical quality of life | Physical function scores were higher in HT users with definite MS (p = .0019) and showed a trend with HT duration (p = .06) |
| [Geng and Tang (2024)](https://pubmed.ncbi.nlm.nih.gov/39073697/#:~:text=Results:%20The%20MR%20analysis%2C%20using,SHBG%20protein%2C%20human) | China researchers used IEU gwas |  | NA | Human | 147,690 | Oestradiol | Female | Incidence | NS associations were found for estradiol |
| [Juutinen et al (2024)](https://pubmed.ncbi.nlm.nih.gov/38442501/) | Finland | MS | 45-54 | Human | 31 | Oestradiol | Female | EDSS, lesion load and whole brain volume | Oestradiol showed a positive correlation with whole brain volume (WBV) on MRI, an inverse correlation with lesion load, serum neurofilament light chain (sNfL), and serum glial fibrillary acidic protein (sGFAP), but no correlation with EDSS |
|  |  |  |  |  |  |  |  |  | ***Key -*** *MS: Multiple Sclerosis. RA: Rheumatoid Arthritis. T1D: Type 1 Diabetes. EDSS :Expanded Disability Status Scale. E1: Oestrone. E2: Oestradiol. E3: Oestriol. EAE: experimental autoimmune encephalomyelitis. Ns: Non significant reported only. OVX: Ovariectomy. VAS: Visual analogue scale. CI: confidence interval. HRT: Hormone replacement therapy. BMD Bone mineral density. HT: Hormone therapy.* |

**Key -** MS: Multiple Sclerosis. RA: Rheumatoid Arthritis. T1D: Type 1 Diabetes. EDSS : Expanded Disability Status Scale. E1: Oestrone. E2: Oestradiol. E3: Oestriol. EAE: experimental autoimmune encephalomyelitis. Ns: ‘Non-significant’ reported only. OVX: Ovariectomy. VAS: Visual analogue scale. CI: confidence interval. HRT: Hormone replacement therapy. BMD Bone mineral density. HT: Hormone therapy.
